# Extended Storage of SARS-CoV2 Nasopharyngeal Swabs Does Not Negatively Impact Results of Molecular-Based Testing

**DOI:** 10.1101/2020.05.16.20104158

**Authors:** Karin A. Skalina, D.Y. Goldstein, Jaffar Sulail, Eunkyu Hahm, Momka Narlieva, Wendy Szymczak, Amy S. Fox

## Abstract

With the global outbreak of the novel coronavirus disease 2019, the demand for testing rapidly increased and quickly exceeded the testing capacities for many laboratories. Clinical tests which receive CE and FDA authorizations cannot always be tested thoroughly in a real-world environment. Here we demonstrate the long-term stability of nasopharyngeal swab specimens for SARS-CoV-2 molecular testing across three assays recently approved by the U.S. FDA under Emergency Use Authorization. This study demonstrates that nasopharyngeal swab specimens can be stored under refrigeration or even ambient conditions for 21 days without clinically impacting the results of the real-time RT-PCR testing.

## Introduction

With the emergence and global spread of the novel coronavirus disease 2019 (COVID-19) caused by the severe acute respiratory syndrome coronavirus 2 (SARS-CoV-2), the development of accurate and timely diagnostic tests has been critical to disease containment. To allow for the extensive testing needs required for effective mitigation strategies, the typical regulatory CE Mark or Food and Drug Administration (FDA) approval processes require a more permissive approach.(1, 2) These streamlined processes may leave individual laboratories and patients at some risk as real-world performance characteristics are assessed within the actual clinical setting. Additionally, the unprecedented rise in necessary testing caused some laboratories to reach maximum testing capacity creating delayed results and the need for prolonged storage of specimens prior to testing.(3) Given the necessity for possible longer-term storage and the lack of robust preclinical data, it is critical to understand the stability of SARS-CoV-2 ribonucleic acid (RNA) following collection of patient nasopharyngeal specimens.

There are limited studies investigating the reliability of viral detection by real time reverse-transcriptase polymerase chain reaction (RT-PCR) over an extended period of time. A study by Druce et al. in 2012 demonstrated that influenza, enterovirus, herpes simplex virus, and adenovirus could be detected by real time RT-PCR reliably for seven days when stored under ambient or refrigerated conditions.(4) Another study published in 2016 by Dare et al. determined that short delays (up to 4 days) in processing influenza nasal and throat swabs did not significantly affect the ability to detect viral particles by real-time RT-PCR.(5) More recently the stability of SARS-CoV-2 detection in different types of storage media over a 14-day period was evaluated.(6) In this study it was found that SARS-CoV-2 was detectable using the Roche Cobas platform over this two-week period regardless of the transport medium.

In our current investigation, we expand upon these previous findings by assessing the detection of the SARS-CoV-2 on multiple commonly utilized Emergency Authorized platforms. Each of these platforms utilizes unique DNA probes with distinct fluorophores as well as differing primer regions within the viral genome. These varied assay constructs may be differentially affected by storage conditions and degradation. Additionally, we assessed viral detection daily over a period of three weeks enabling extensive precision data to be produced and analyzed for each platform providing essential laboratory performance characteristics in an actual clinical laboratory environment.

## Methods

### Creating a Shared Pool

Thirty anonymized remnant nasopharyngeal swab samples collected in viral transport media (VTM) which were positive for SARS-CoV2 in our laboratory were combined to create a large pool of positive patient-based material. Given the abundant viable virus present in many of these clinical samples, the decision was made to heat-inactivate the large pooled material at 56°C as previously reported.(7) A total of 126 equal aliquots of the pooled, inactivated material were then stored in one of 2 maintenance conditions, either ambient temperature (18-25°C) or within a refrigerator maintained between 2-8°C. Preparation and initial storage of the pooled material occurred on day 0 and testing began the following day.

### Instrumentation

This study utilized three different automated real-time reverse-transcriptase polymerase chain reaction (RT-PCR) in vitro diagnostic platforms (Luminex ARIES, Panther Fusion, and Abbott m2000) currently in use for clinical testing of SARS-CoV-2 at the Department of Pathology, Division of Virology, Montefiore Medical Center, Bronx, NY. All technicians who performed testing were trained in the operation and maintenance of these instruments. All three platforms have been approved for use by the United States Food and Drug Administration under the Emergency Use Authorization (EUA).

Each platform’s PCR reaction is designed to amplify 2 target regions within the SARS-CoV-2 genome (Luminex: ORF1a/N, Abbott: RdRp/N and Hologic: separate ORF1a regions).(8-10) However, the Hologic Panther and Abbott m2000 utilize the same fluorescent channel for reporting of the amplification product thereby only providing a single Ct for the merged fluorescent intensity. The Luminex ARIES platform utilizes two fluorophores for each amplification probe allowing for individual detection of the amplicons and providing separate Ct values for ORF1a and N. Additionally, whereas the internal amplification control (IC) utilized by the ARIES amplifies human ribonuclease P (Rnase P) within the collected sample, both the Panther Fusion and the m2000 utilize a spiked exogenous amplification control during sample preparation and would thus not be subject to the effects of long term storage. All materials were run according to manufacturers’ instructions for use by trained licensed personnel.(8-10)

### Analysis

The cycle thresholds (Ct) for the amplification of each platforms target(s) and IC were recorded and graphed using GraphPad Prism software. Additional statistical analysis of the recorded data, including linear regression, correlation, coefficient of variation and paired *t* tests were performed using Microsoft Excel.

## Results

### Shared Pool Samples

Over the course of the 21 days, the pooled materials were tested alongside clinical samples and the cycle thresholds (Ct) recorded for each platform at both ambient (Figure 1) and refrigeration temperatures (Figure 2). A total of 102 aliquots were ultimately run for a total of 244 data points. Qualitative detection of the virus was seen across all conditions and platforms demonstrating 100% accuracy with the intended result regardless of sample age or storage condition. While the difference between the means of Ct at the two storage conditions were statistically significant for the Fusion (p<0.0001) and m2000 (p=0.037), there was no statistically significant difference in either target’s Ct on the ARIES (N p=0.26, ORF p=0.72) (Table 1). Despite statistical significance, the difference in mean Ct values from the two storage conditions would not be clinically significant. The largest mean Ct deviations between the two temperatures occurred on the m2000 (1.5 cycles), however, a Ct change of this magnitude would only be expected to impact samples with exceedingly low viral burdens and only have a minimal impact in the clinical testing environment.

**Figure 1.**
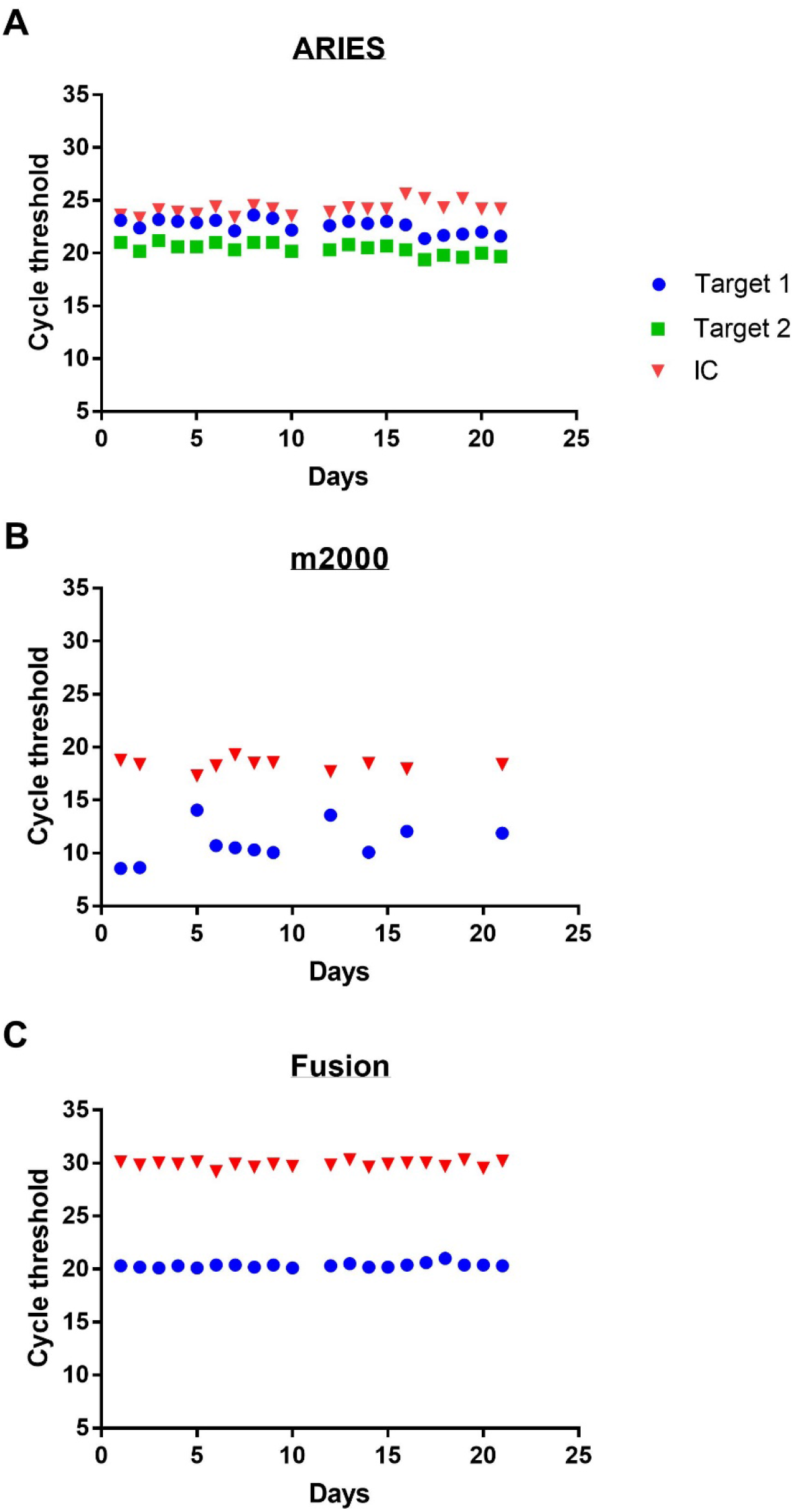
Cycle Threshold of SARS-CoV2 in VTM stored at ambient temperature over time on 3 automated real time RT-PCR machines Luminex ARIES (A), Abbott m2000 (B), and Hologic Panther Fusion (C) Target 1 refers to the N target for the ARIES and both targets for the m2000 and Fusion. Target 2 refers to the ORF target for the ARIES.

**Figure 2.**
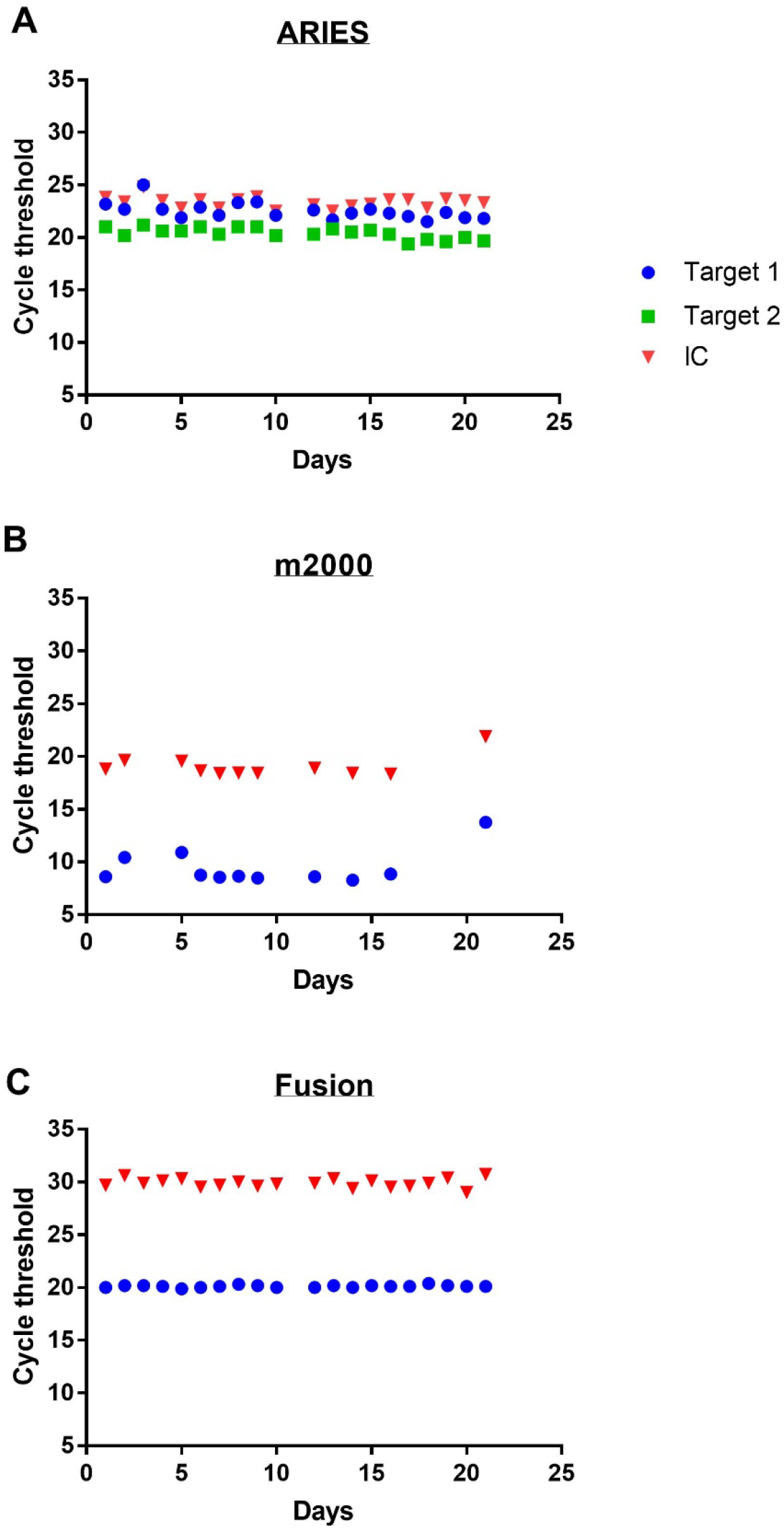
Cycle Threshold of SARS-CoV2 in VTM stored under refrigeration over time on 3 automated real time RT-PCR machines Luminex ARIES (A), Abbott m2000 (B), and Hologic Panther Fusion (C) Target 1 refers to the N target for the ARIES and both targets for the m2000 and Fusion. Target 2 refers to the ORF target for the ARIES.

**Table 1.** Comparison of Storage Temperature on the Average Ct. Significance was calculated using a paired t-test.

|  |  | Mean | SD |  |
| --- | --- | --- | --- | --- |
| Fusion | 20°C | 20.34 | 0.2062 | n=20 |
|  | 4°C | 20.12 | 0.1196 | n=20 |
|  | Difference | -0.22 | 0.1795 | p<0.0001 |
| m2000 | 20°C | 10.96 | 1.793 | n=11 |
|  | 4°C | 9.45 | 1.658 | n=11 |
|  | Difference | -1.506 | 2.077 | p=0.037 |
| ARIES<br>N Target | 20°C | 20.41 | 0.522 | n=20 |
|  | 4°C | 20.26 | 0.7687 | n=20 |
|  | Difference | -0.15 | 0.5835 | p=0.2646 |
| ARIES<br>ORF<br>Target | 20°C | 22.58 | 0.6348 | n=20 |
|  | 4°C | 22.53 | 0.794 | n=20 |
|  | Difference | -0.05 | 0.6295 | p=0.7264 |

Overall consistency of Ct values over the study period, for all assays and storage conditions was very good with the Fusion demonstrating slightly tighter grouping over time with the coefficients of variation (CV) of only 0.59% and 1.01% for refrigeration and room temperature respectively. The slopes of the linear regression through each platform/temperature pair ranged from -0.075 (ARIES 4°C) to 0.129 (m2000 RT) indicating only minimal deviation from the horizontal for all platforms over time. The strengths of these correlations over time as represented by the R^2^ also demonstrate near *0* values (Table 2) indicating very little impact over time.

**Table 2.**
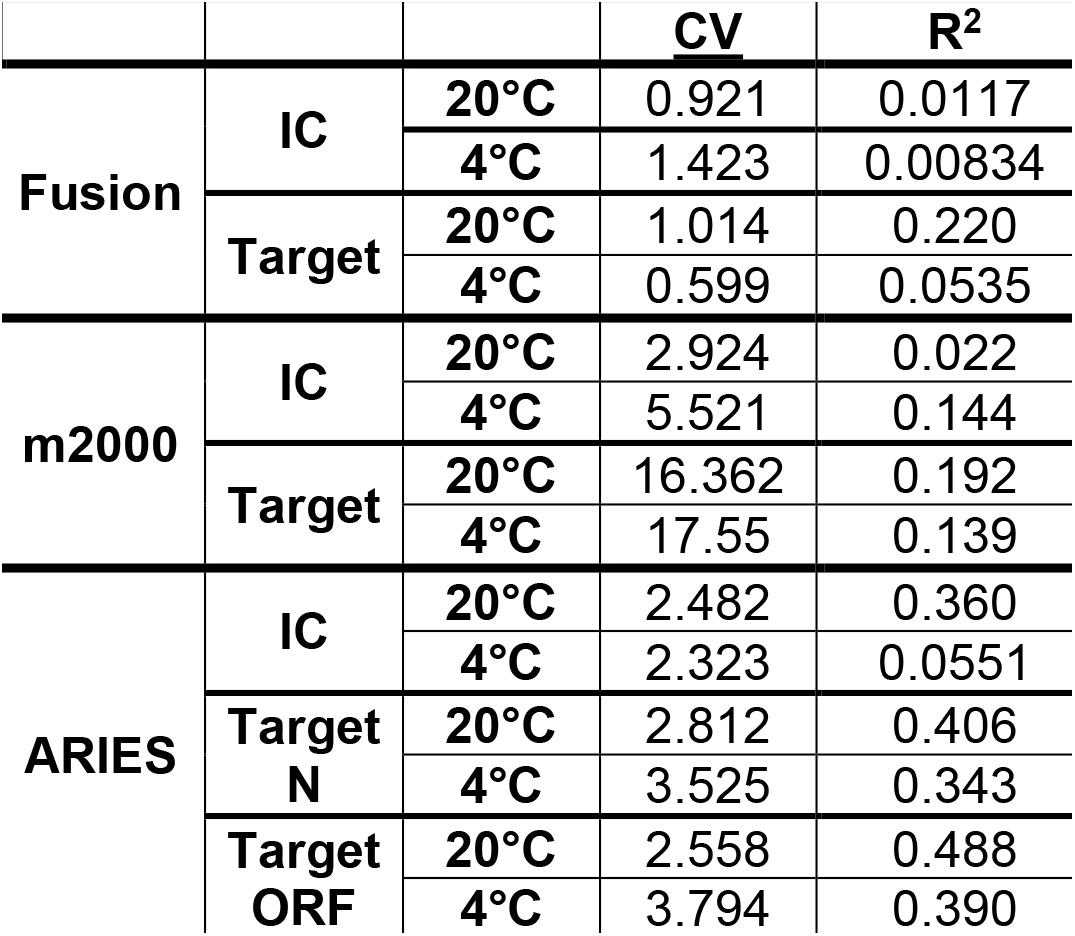
Statistical Analysis of the Cycle Thresholds (Ct)

### Patient Samples

Given the ideal conditions associated with the pooled material and the possibility of RNAse degradation through the heat inactivation step, we next sought to confirm the integrity of our findings using actual stored clinical samples. Seven positive clinical samples which had been held at 4°C were repeated on day 35 after collection and run on the Hologic Panther. These unadulterated samples included some containing abundant mucus which would be expected to contain RNA degrading RNAses. All 7 samples remained positive when performed at the later time point. The Ct values for these specimens ranged from 13.2 to 30.6.

## Discussion

Testing delays and laboratory backlogs have contributed to the worsening spread of COVID-19 infections. While the testing situation in the United Kingdom and United States is improving, the number of tests performed per 100,000 individuals still lags behind other developed countries such as Demark and Italy.(11) In an effort to broaden testing capabilities, the US FDA allows some manufacturers to market their assays under the less strenuous review process of Emergency Use Authorization (EUA). It has been demonstrated that for some platforms that a manufacturers’ claims provided through this more liberal review pathway may significantly overestimate performance characteristics in a true clinical environment.(12)

For these reasons we sought to evaluate the performance of three separate EUA platforms over a lengthy period of time. Our data supports others’ findings that viral RNA may remain stable for molecular testing for some time.(6, 12, 13) All of the tested platforms, each designed to amplify completely different PCR amplicons, performed extremely well, providing precise Ct values throughout the testing period. Most surprisingly, there was only a very minimal, and clinically insignificant, impact of storage condition on the Ct values for all assays. Storage of pooled material at ambient temperature over this extended time course did not alter the test results. This demonstrated stability may allow laboratories with limited cold storage capabilities to maintain specimens at ambient temperatures without an impact to clinical performance.

Finally, we continued to add to previous studies by demonstrating that even samples with abundant mucoid material, whose composite white blood cell content could hamper assay performance and degrade RNA, remained positive at 35 days.

While the efforts of manufacturers to produce, and governments to obtain, testing for a global population is admirable, even broader based test access will be required to further mitigation and containment strategies. Public policy allowing for easier test approval must be balanced against ensuring the safety and efficacy of that testing. With the continued spread of the SARS-CoV-2 virus, globally, clinical laboratories will remain a vital resource in evaluating performance characteristics under real world conditions.

## Data Availability

All data will be made available upon request.

## Notes

### Competing Interest Statement

The authors have declared no competing interest.

### Funding Statement

No external funding was used for this project.

